# Enfortumab vedotin-induced cutaneous toxicities and their association with survival in urothelial carcinoma

**DOI:** 10.64898/2026.05.19.26353579

**Authors:** Eudora Lee, Ralina Karagenova, Charles Lu, Parisa Farokh, Marjan Azin, Federico Repetto, Soma Jobbagy, Rosalynn M. Nazarian, Kerry Reynolds, Shadmehr Demehri, Philip J. Saylor, Lirit Fuksman, Yevgeniy R. Semenov

## Abstract

**Key Points:** *Question:* Among patients with advanced or metastatic urothelial cancer treated with enfortumab vedotin (EV), is development of EV-induced cutaneous adverse events (cAEs) associated with improved survival, and how does this association compare with cutaneous immune-related adverse events (cirAEs)?

*Findings:* In a multi-institutional retrospective cohort study, EV-induced cAEs were associated with significantly progression-free survival (hazard ratio, 0.60; 95% CI 0.31-0.67, p<0.001) and overall survival (hazard ratio, 0.46; 95% CI 0.31-0.67, p<0.001) across landmark analyses.

*Meaning:* EV-induced cAEs are independently associated with improved survival in patients, suggesting potential prognostic value during treatment of advanced urothelial cancer.

**Importance:** Enfortumab vedotin (EV) is an antibody-drug conjugate approved for the treatment of locally advanced or metastatic urothelial cancer (la/mUC). Cutaneous adverse events (cAEs) are common during EV therapy, with prior studies suggesting an association between EV-related cAEs and improved survival; however, there is insufficient data to delineate the survival benefit of EV-induced cAEs from those associated with concurrent immune checkpoint inhibitors (ICIs).

**Objective:** This study aims to evaluate the association of EV-induced cAEs and survival, and to characterize the timing and morphology of EV-induced cAEs.

**Design:** We conducted a multi-institutional retrospective study of patients with la/mUC treated with EV between 2020 and 2025.

**Setting:** Multicenter academic referral center.

**Participants:** A total of 449 EV-treated patients were included. Patient characteristics were extracted manually, and likelihood scoring was used to attribute cAEs to either EV or other etiologies.

**Exposure:** EV treatment.

**Main Outcomes and Measures:** We estimated progression-free (PFS) and overall (OS) survival using Kaplan-Meier method. Multivariable time-varying and landmark Cox regression models were used to evaluate associations between EV-induced cAE and survival. Sensitivity analyses were performed at landmarks from 15 to 105 days.

**Results:** Of 449 patients, 206 (45.9%) developed a cAE; 39 (18.9%) were high-grade and 127 (61.7%) were attributed to EV. The most common cAEs were pruritus (41.3%), unspecified and desquamating dermatitis (37.3%), and morbilliform dermatitis (27.7%). Across all treatment groups, survival was longer in patients with EV-induced cAEs. Developing an EV-induced cAE was protective across all examined landmark times, with hazard ratio (HR) 0.60 (95% CI: 0.43-0.82, p<0.001) for PFS and HR 0.46 (95% CI: 0.31-0.67, p<0.001) for OS at primary landmark time of 30 days. Early-onset EV-induced cAEs were protective at all landmark times and high-grade EV-induced cAEs were not associated with worse survival.

**Conclusions and Relevance:** EV-induced cAEs were independently associated with improved PFS and OS in patients with la/mUC, even after accounting for immortal time bias and ICI exposure. Distinguishing EV-induced cAEs from other etiologies in timeline and morphology may help guide oncology and dermatology management.

## Introduction

Enfortumab vedotin (EV) is an antibody-drug conjugate (ADC) that has rapidly emerged first as monotherapy then in combination with pembrolizumab (an immune checkpoint inhibitor (ICI)) as the first-line systemic treatment of choice for locally advanced or metastatic urothelial cancer (la/mUC)^1,2^. EV consists of monomethyl auristatin E, a microtubule-disrupting agent, linked to an anti-nectin-4 monoclonal antibody. Nectin-4 is highly expressed in urothelial cancer cells and epidermal keratinocytes^3^. Beyond direct cytotoxicity, EV also induces immunogenic cell death, supporting its combination with ICIs^4^. However, cutaneous adverse events (cAEs) have been reported in up to 45% of patients and represent the most common high-grade toxicity associated with EV in clinical trials^2^. It is unknown whether EV-induced cAEs are prognostically meaningful, which is complicated by the frequent co-administration of EV with ICI where cAEs have been associated with improved survival.

While several observational cohorts have examined the association between EV-induced cAEs and patient outcomes, these were limited by small sample sizes^5–8^, reliance on International Classification of Diseases (ICD) codes rather than clinician-assessed cAE ascertainment^7^, inability to differentiate EV-from ICI-induced cAEs, and conflicting results. Some of those analyses suggested a survival benefit^6,7^ and others found no survival benefit associated with these toxicities^8,9^.

Furthermore, limited data exists differentiating between EV-versus ICI-driven etiologies. Prior studies have established a survival benefit to cutaneous immune-related adverse events (cirAEs), including urothelial carcinoma^10–12^. Thus, there remains a challenge to differentiate whether the captured survival benefit represents a true, independent effect of EV or from concurrent pembrolizumab administration. Data characterizing morphology and timing of EV-induced cAEs, which may aid causal attribution and inform clinical management, remain limited.

This study leverages a large multi-institutional cohort to examine the epidemiology, morphology, timing, management, and survival outcomes associated with EV-induced cAEs and provides a framework to differentiate these from ICI-induced cAEs. In doing so, this study provides a roadmap to diagnosing and managing these cutaneous toxicities in a rapidly growing patient population.

## Methods

We conducted a multi-institutional retrospective cohort study of 449 la/mUC patients treated with EV between 2020 to 2025 at the Mass General Brigham and the Dana-Farber Cancer Institute (MGBD). Patient demographics were extracted from the MGBD Research Patient Data Registry and patients who did not receive EV or who did not have stage 3 or 4 urothelial cancer were excluded. Manual chart review was conducted to extract AE characteristics, including cAE status, dates of onset and resolution of first and most severe cAE, morphology, and severity. Cancer treatment variables included start and end dates of EV treatment, initial EV dose, prior cancer treatments, treatment group (EV-only, EV with pembrolizumab (EV+ICI), or EV with any other non-ICI agent (EV+other)), and tumor response. Management of cAE variables included evaluation by dermatology (yes/no), topical corticosteroid use (yes/no), systemic corticosteroid use (yes/no, dose and duration), biologic therapy use (yes/no). Patient outcomes included vital status, cause of death, date of death, date of progression, and date of last contact.

Ascertainment of cAE status was conducted by two trained independent research analysists (E.L. and R.K.) in accordance with a standardized method developed and previously validated by our group^10,13,14^, with arbitration of discordant cases by a third reviewer (Y.R.S., board-certified dermatologist with expertise in oncodermatology). Briefly, a likelihood score between zero and four was assigned to each cAE, where 0 represented a pre-existing eruption prior to EV initiation, 1 represented an eruption that is highly unlikely to be secondary to EV, through 4 representing an eruption that is highly likely to be secondary to EV. Scores were determined by manual review of patient charts and clinician documentation, including photographs, morphology, timing, course of the eruption, and histology, where available. A detailed explanation of this methodology is presented in the Supplement Supplementary Table 1^15–17^. Likelihood scores of 3 or 4 were considered EV-induced and scores of 0-2 considered non-EV-induced. If there was no cAE, likelihood scoring was not applied.

Morphology was documented based on clinical notes (if the patient was evaluated by dermatology), or where not available, by analyst assessment of clinical descriptions and photographs. Histopathologic findings, when available, were used to support this classification. Common Terminology Criteria for Adverse Events (CTCAE) version 5.0^18^ was used to grade cAE severity. Although the majority of patients did not receive EV as part of a clinical trial, radiologic response was assessed using clinical documentation and the Response Evaluation Criteria in Solid Tumors (RECIST) criteria as best as possible, acknowledging that follow-up timing for restaging was variable for assessing progression-free survival (PFS) and overall survival (OS) was the primary outcome of interest^19^. If multiple cAEs developed, analyses were performed for the most severe cAE. Outcomes were compared to a control group that did not develop cAEs.

PFS and OS were estimated using the Kaplan-Meier method, and survival curves between treatment types were compared using log-rank test. To account for immortal time bias, landmark analysis with multivariable Cox proportional hazards regression (CoxPH) models with primary landmark time of 30 days were adjusted for sex, race, ethnicity, age at EV initiation, treatment group, National Cancer Institute Comorbidity (NCI) Index, systemic corticosteroid use, cumulative number of EV infusions, and cancer stage at EV initiation.

As an alternative modeling approach, time-varying CoxPH treating EV-induced cAE as time dependent covariate and adjusting for the same variables was used. Cumulative number of EV infusions exhibited a skewed distribution with few patients receiving more than 50 infusions (Supplementary Table 2). Therefore, the variable was capped at its 95th percentile and included in the model as a restricted cubic spline term with nonlinear effect which is not interpreted in the analysis. Cutaneous AE grade was excluded from the model given its collinearity with likelihood score (p=0.02). Finally, landmark CoxPH analyses compared the survival benefit of early-onset cAEs (occurring by 15 days after EV initiation) versus late-onset cAEs (15 days after EV initiation but before landmark time).

Sensitivity analysis was performed by additionally adjusting for initial EV dose, Eastern Cooperative Oncology Group Performance Status (ECOG-PS), and prior ICI and chemotherapy exposure. Pairwise comparisons of the median time to worst cAE onset between treatment groups were performed using Wilcoxon rank sum test with Benjamini-Hochberg correction. Alpha of 0.05 was used as the significance threshold. Statistical analyses were conducted in R v4.5.0.

## Results

Of 449 patients, 172 (38%) were treated with EV only, 256 (57.0%) were treated with EV and pembrolizumab (EV+ICI), and 21 (4.7%) were treated with EV and sacituzumab govitecan (ADC), ALX148 (anti-CD47), or GEN-009 vaccine (EV+other). At time of chart review, 59.9% of the total cohort was deceased, with 91.8% of deaths attributed to cancer. Baseline characteristics of the cohort are presented in Table 1 and cAE treatment characteristics by severity are presented in Supplementary Table 3. There was no significant difference in cAE frequency when comparing by prior ICI or chemotherapy exposure (Supplementary Table 2). The distribution of total EV infusions and initial EV dose differed significantly between patients with and without cAEs. Cutaneous AEs were present in 206 (45.9%), of which 39 (18.9%) were high-grade (3 or above). The most common cAEs were pruritus (41.3%), followed by unspecified eruption, which include rash (unspecified) and desquamating dermatitis (37.3%), and morbilliform dermatitis (27.7%) (Table 2). Supplementary Table 4 shows morphology frequencies stratified by likelihood score within the treatment groups. Of note, unspecified eruptions were defined as those lacking sufficient narrative or photographic documentation to allow adjudication. A total of 84 (40.8%) patients with cAEs were evaluated by dermatology. Among all cAEs, 127 (61.7%) were attributed to EV. The most common non-cutaneous AEs were neuropathy (37.9%), fatigue (18.9%), and diarrhea (8.9%) (Supplementary Table 6). Neuropathy has been reported to be independently associated with improved OS^9^. Neuropathy distribution did not differ significantly by treatment group or cAE status, reducing concern for confounding.

**Table 1.**
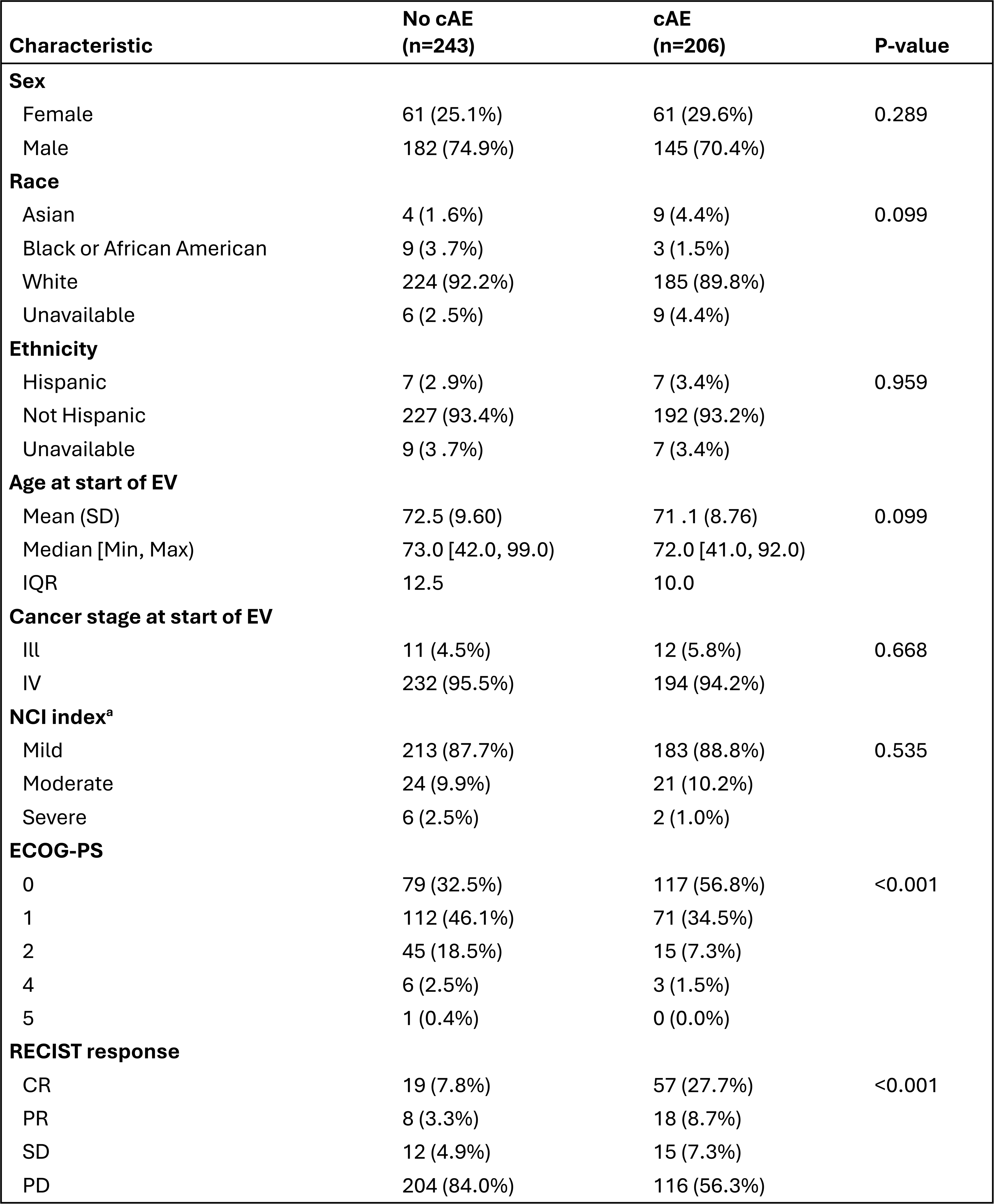
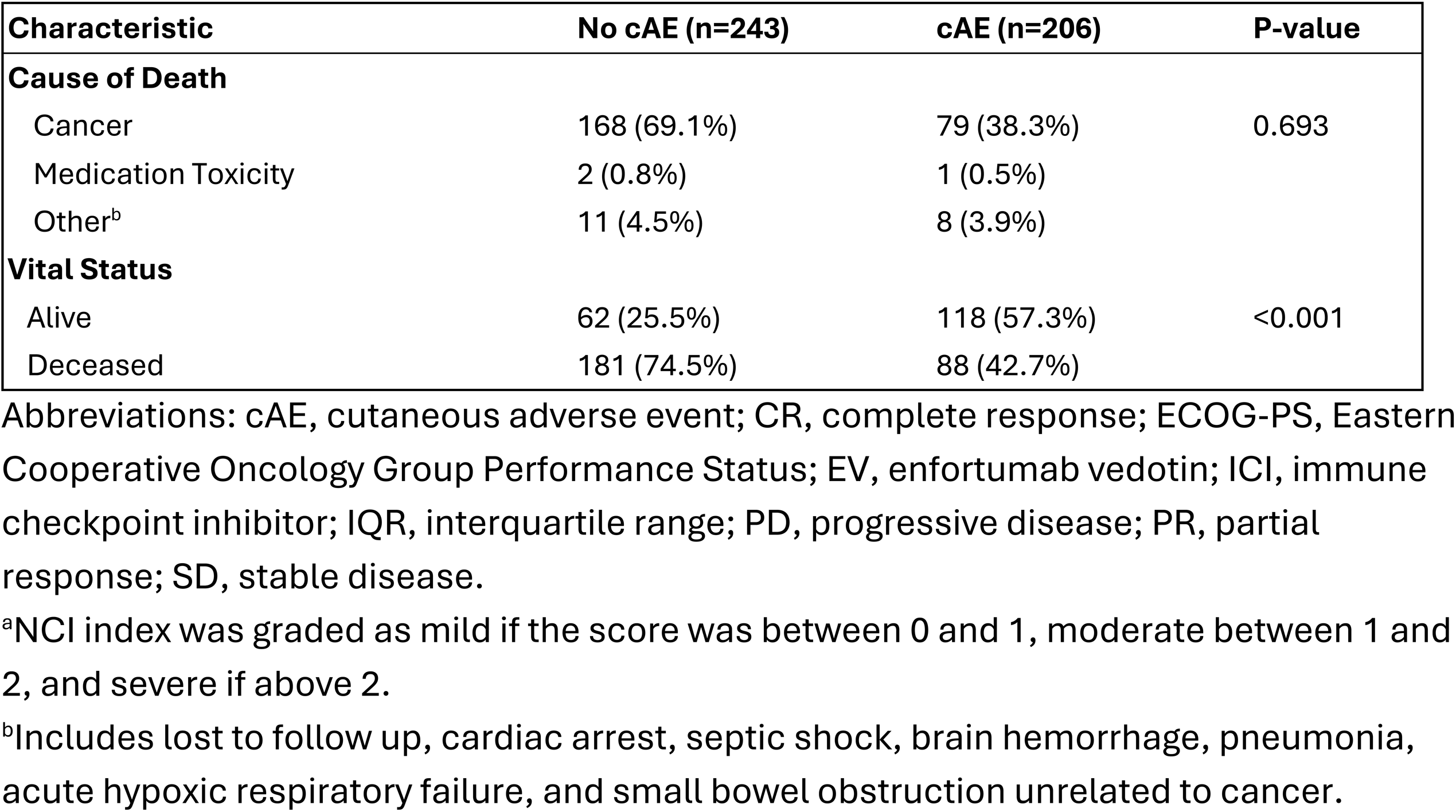
Demographic Characteristics, Treatment exposure, and Clinical Outcomes Stratified by Development of Cutaneous Adverse Event.

**Table 2.**
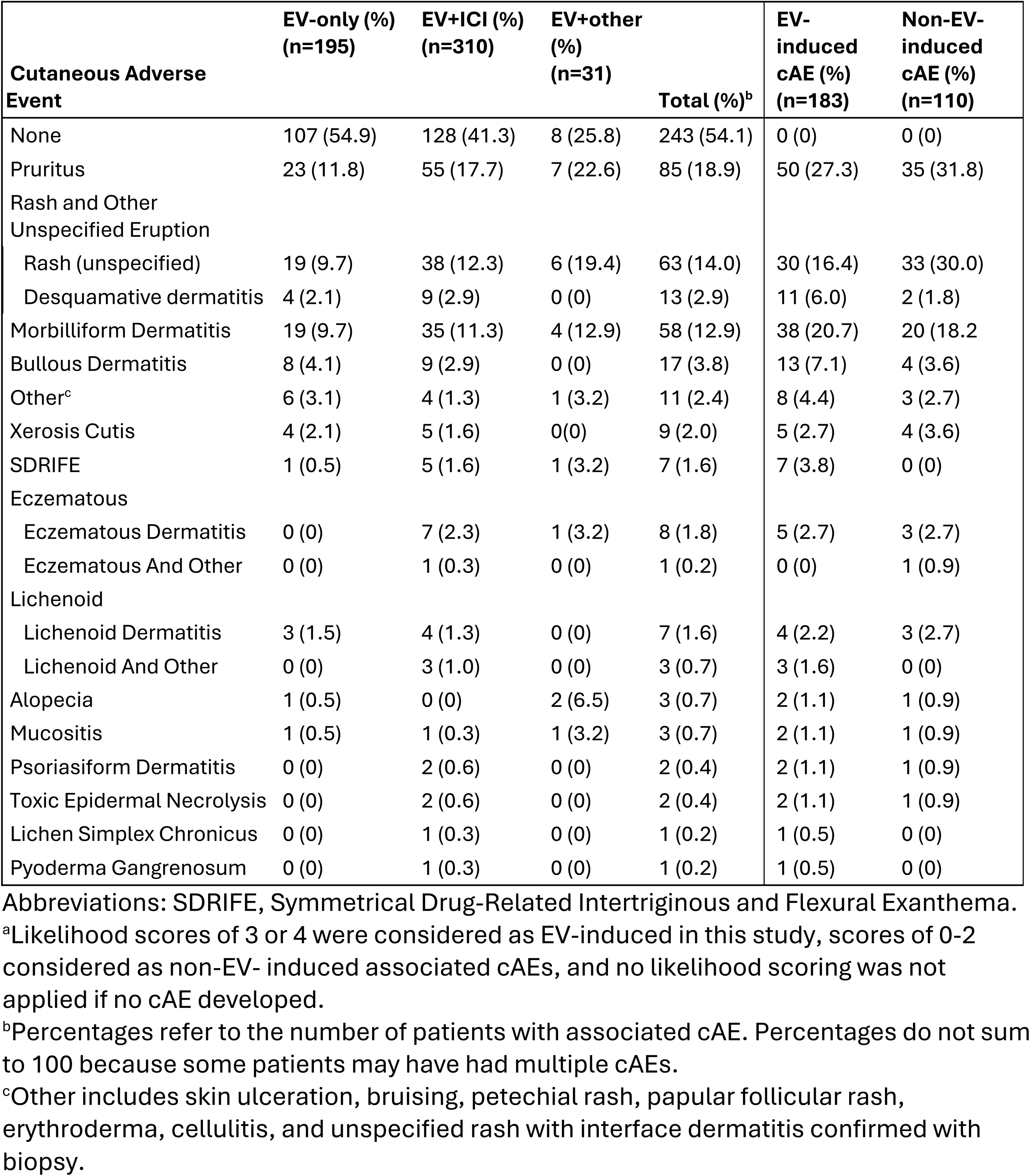
Cutaneous Adverse Event Morphology Distribution Stratified by Treatment Group and Likelihood Scoring^a^.

Median time from first EV infusion to onset of worst cAE was 17 days, and 129 (62.6%) patients developed their worst cAE by day 30 (Supplementary Figure 1). There was no significant difference between EV-only group and EV+ICI group (15 vs 21 days, p=0.40). However, EV+other group had significantly earlier eruptions than EV+ICI (8 vs 21 days, p=0.006) or EV-only (8 vs 15 days, p=0.006), though the EV+other group had small cohort size (n=13) (Table 3). Median time to cAE resolution was 48 days, with no significant difference among the three treatment groups. Median time to onset was also similar across cAE morphologies, ranging from 13 to 22 days, except for lichenoid eruptions which had the highest median of 49 days (Figure 1). By reverse Kaplan-Meier estimator, the median follow-up time was 21.6 months.

**Figure 1.**
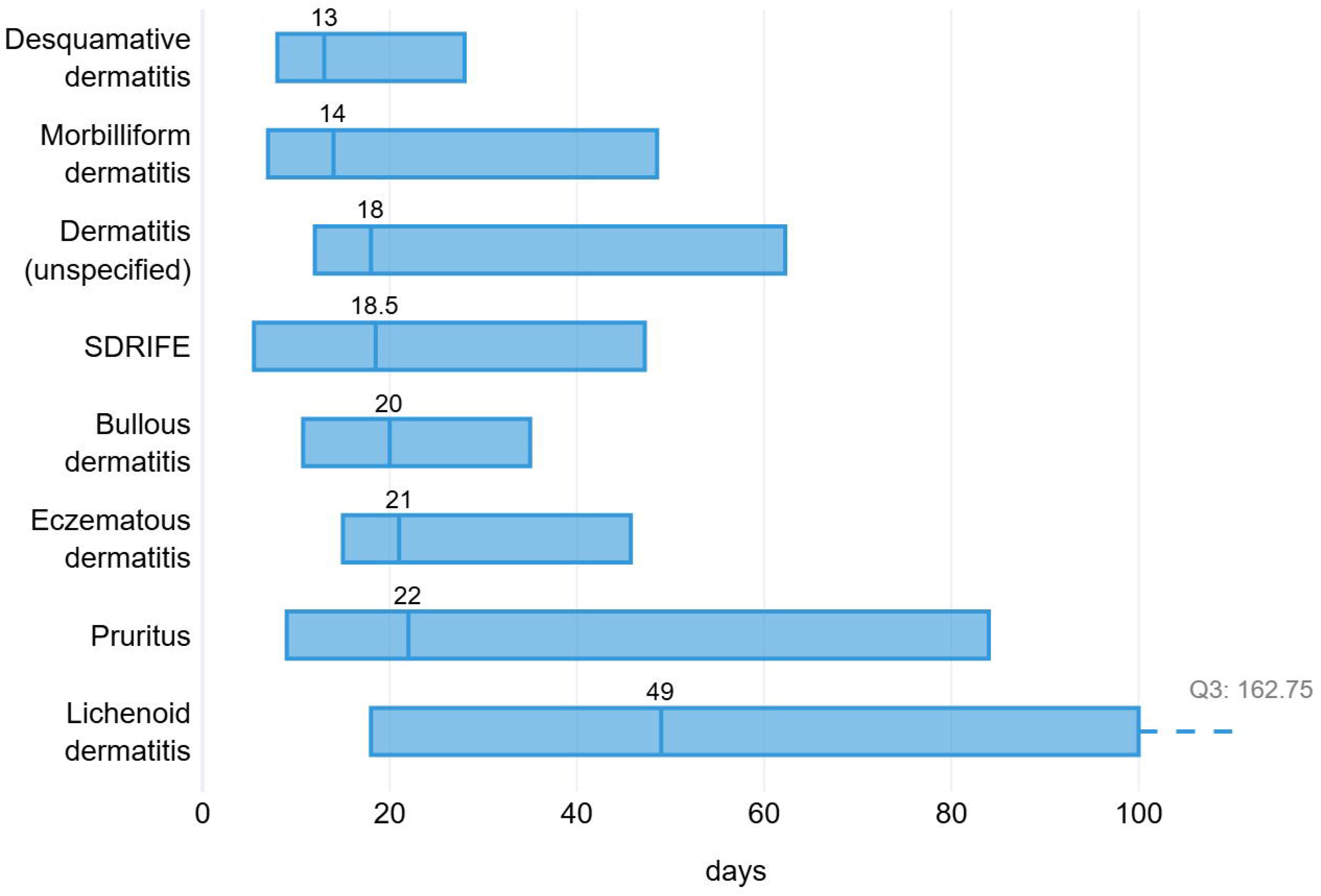
Temporal distribution of cutaneous adverse events (cAE) morphologies. The figure illustrates the relative timing and overlap of distinct cAE morphologies, highlighting early- and late-onset patterns.

**Table 3.**
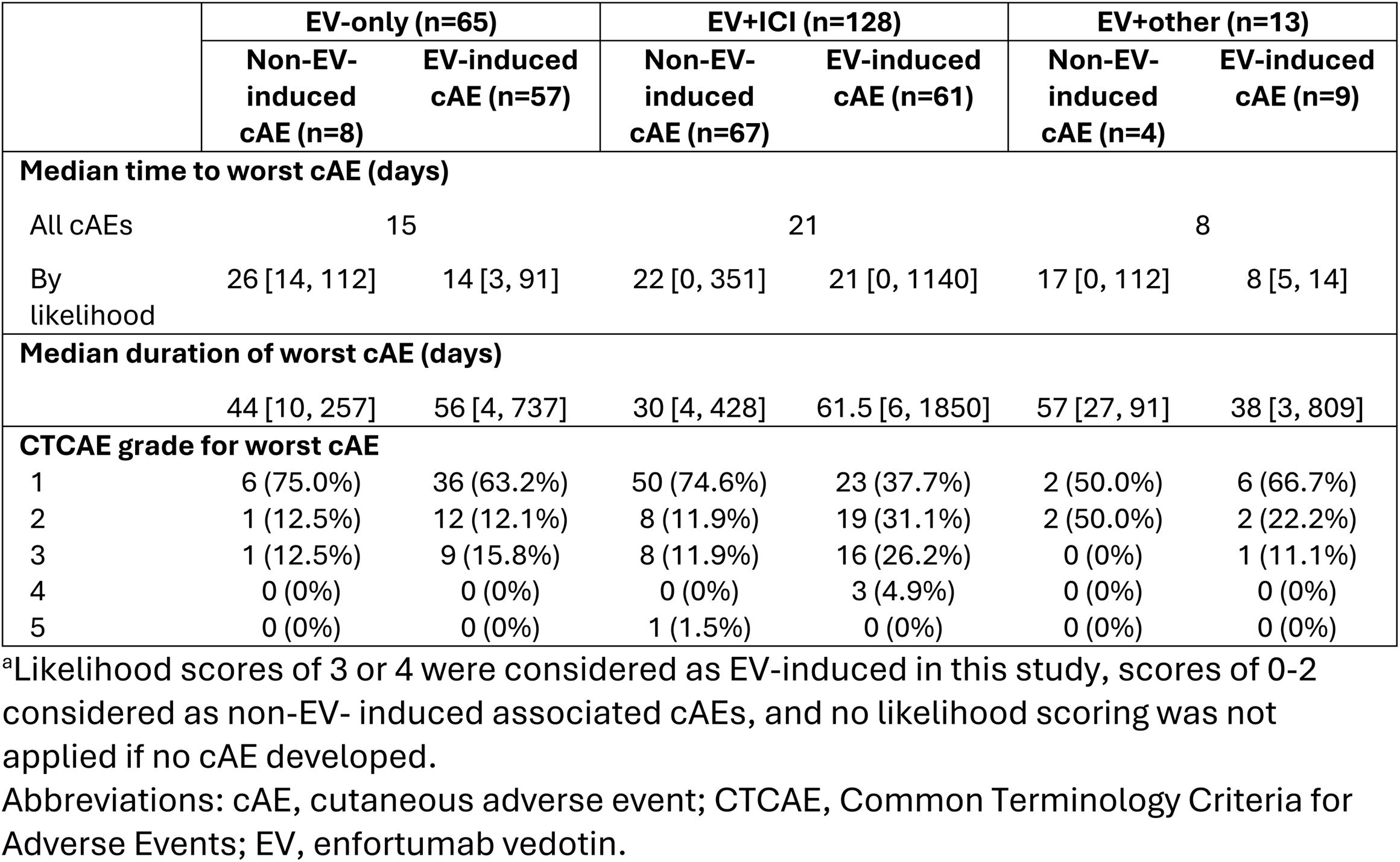
Timing and Severity of cAEs Stratified by Treatment Group and Likelihood Scoring^a^.

Overall, most cAEs were low-grade (64.6%, 57.0%, 61.5% for the EV-only, EV+ICI, EV+other groups, respectively, p=0.814). Between EV-only and EV+ICI cohorts, there was no significant difference in severe cAEs (p=0.14). However, EV+ICI group had higher odds of developing any cAE compared to EV-only (OR=1.64, 95% CI 1.09-2.49, Fischer’s exact test p = 0.01), but lower odds of that cAE being EV-induced (OR=0.63, 95% CI 0.40-0.99, Fisher’s exact test p=0.04). Among patients with cAEs, 80.1% were treated with topical corticosteroids, out of whom 18.2% additionally received a systemic corticosteroid, 3.0% received a systemic biologic (dupilumab), and 1.8% received all three. The median duration of systemic corticosteroids was 34 days. The median cumulative dose of systemic corticosteroids was higher, though not reaching statistical significance, among patients with high-grade eruptions (p=0.687). Distribution of management by severity of cAEs is shown in Supplementary Table 3. Among those that developed a cAE, 21 out of 29 with available histopathology results were considered EV-induced. Of the patients with available histopathology that were treated with dupilumab, 3 were EV-induced and 4 were non-EV-induced. The most common histopathologic descriptors in patients who received dupilumab was drug hypersensitivity (57.1%), followed equally by eczematous and lichenoid patterns (28.6%).

For both PFS and OS in EV-only group, patients with EV-induced cAEs had significantly higher survival compared to those not developing a cAE or developing a non-EV-induced cAE. In EV+ICI group, patients with any cAE had significantly higher survival than those without cAE (Figure 2). Results for EV+other group can be found in Supplementary Figure 2 due to smaller sample size.

**Figure 2.**
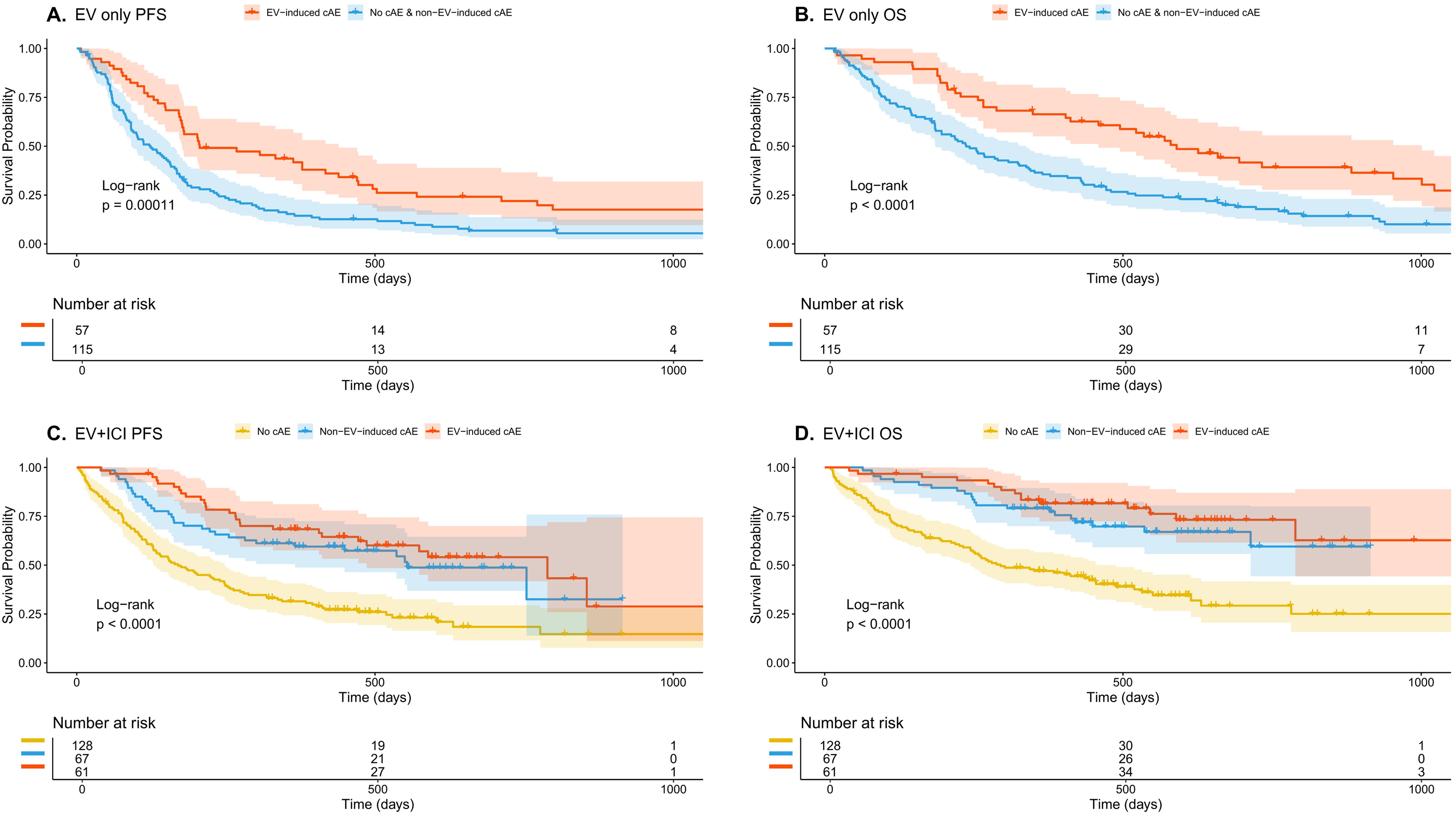
Kaplan-Meier Estimates of Progression-Free Survival (PFS) in EV-only (panel A) and EV+ICI (panel B) Treatment Groups and Overall Survival (OS) in EV-only (panel C) versus EV+ICI (panel D), stratified by EV-induced, non-EV-induced cAE, or no cAE

In the EV-only group, median PFS was 5.2 months and median OS was 10.9 months. In EV+ICI group, the PFS and OS was 10.6 and 21 months, respectively. Median survival times for each treatment group stratified by cAE status are summarized in Supplementary Table 5.

In landmark CoxPH analysis, EV-induced cAEs were independently associated with improved PFS (HR 0.60, 95% CI 0.43–0.82, p<0.001) and OS (HR 0.46, 95% CI 0.31–0.67, p<0.001). The protective association of EV-induced cAEs remained consistent across multiple landmark points (Supplementary Figure 3A and 3B). In time-varying CoxPH analysis, this effect was similar, with EV-induced cAEs showing protective effect for PFS (HR 0.56, 95% CI 0.42–0.74, p<0.001) and OS (HR 0.51, 95% CI 0.37–0.70, p<0.001). Additional sensitivity analysis is shown in Supplementary Figure 4, and although the effect sizes changed slightly numerically, the overall conclusions remained unchanged.

Non-EV-induced cAEs were not significantly associated with either outcome in both models. However, they were protective for OS (HR 0.55, 95% CI 0.33–0.92, p=0.02) in the EV+ICI subgroup, likely reflecting cirAE benefit. The discrepancy between the non-significant interaction in the full cohort contrasted with the significant effect in the EV+ICI subgroup likely reflects heterogeneity from inclusion of EV-only patients, attenuating the effect.

When stratifying EV-induced cAEs by early (≤15 days) versus late-onset, only early-onset conferred a protective signal for PFS (HR 0.57, 95% CI 0.40–0.82, p=0.002) and OS (HR 0.42, 95% CI 0.28–0.64, p<0.001). The result held across all examined time points (Supplementary Figure 3C and 3D). Severe EV-induced cAEs were not associated with significantly worse PFS or OS across all landmark times.

## Discussion

This large multi-institutional cohort demonstrates that EV-induced cAEs are common and prognostically meaningful, particularly when occurring within the first 15 days of EV initiation. Although emerging studies have shown some benefit ^6–9^, the prognostic significance of EV-induced cAEs in patients with la/mUC remains incompletely defined. Here, we build upon a growing but mixed–body of literature by examining survival differences in EV-driven cAEs separately from ICI-driven etiologies.

This study included 449 patients from the MGBD, with 38% treated with EV alone and the remaining receiving EV with either ICI (pembrolizumab) or a non-ICI agent. Nearly half of the cohort (45.9%) developed a cutaneous toxicity with 28.3% experiencing EV-induced cAEs, reflecting previous evidence from clinical trials and real-world data^2,6,7^, with 19% developing a severe, grade ≥3 event.

Our findings suggest that EV-driven cAEs displayed prognostic benefit, with early cAEs conferring a stronger protective signal across both landmark and time-varying analyses, compared to patients who did not develop a cAE. In those treated with EV plus pembrolizumab, developing any cAE (attributed to either agent) was also more protective than not developing a cAE. As supported by existing literature, EV-induced cAEs occurred early in the treatment course, with a median onset of 17 days following EV initiation and almost two-thirds of patients reached peak cAE severity by one month of treatment. The median onset of non-EV-induced cAEs in the EV plus pembrolizumab group had median onset of 22 days, which is earlier than cirAE onset reported in literature, typically occurring 4-6 weeks following initiation of treatment in clinical trials^20^ and 16 weeks in real-world observational data^15^. This suggests that the presence of EV may drive earlier cirAE presentation. Interestingly, lichenoid eruptions had a higher median time to onset of 7 weeks as compared to other morphologies. These temporal distinctions may be of value for both oncologists and dermatologists when determining the most likely agent responsible for an eruption, as this may directly influence toxicity management and decisions regarding treatment interruption or modification. Most patients who developed a cAE were treated with a topical corticosteroid, with fewer than one-fifth of patients receiving a systemic corticosteroid. Dupilumab was used in cases with histopathologically confirmed eczematous dermatitis or presence of eosinophils, regardless of EV versus ICI attribution, suggesting clinicians tailored biologic therapy to histologic pattern rather than causative agent.

Due to small sample size and variety in cAE type, it is challenging to identify a consistent, histologic feature of cytotoxicity that may be more associated with EV compared to an inflammatory pattern that may be associated with a cirAE. However, previous work characterizing the histopathologic patterns of EV-induced cAEs found that all cases showed interface dermatitis with one of four patterns: classic vacuolar, cytotoxic with epidermal dysmaturation, necrolytic SJS/TEN-like, or spongiotic interface^21^. Our findings were consistent with this prior work, with histopathology from EV-induced cAEs showing interface dermatitis with necrolysis, eosinophils, spongiotic interface, vacuolar interface, which are consistent with findings of cytotoxic interface dermatitis. Future studies that build on these histologic patterns may further delineate between EV-induced cAEs and other etiologies.

Moreover, our study found that severe EV-induced cAEs do not portend a worse prognosis. This is in contrast to prior analyses using post-marketing surveillance data from the U.S. Food and Drug Administration Adverse Event Reporting System that identified increased mortality associated with rare, severe cAEs related to EV, including Stevens-Johnson Syndrome and toxic epidermal necrolysis^22^. However, this data is restricted by self-reporting, favoring an overrepresentation of severe events. This, along with the lack of a comparator group, limits causal inferences. In contrast, our cohort-based study represents real-world data with time-to-event analyses, enabling a more robust evaluation of EV-driven cAEs and associated outcomes.

The mechanistic basis for EV-induced cAEs may reflect on-target nectin-4 expression in tumor cells and epidermal keratinocytes, potentially serving as a pharmacodynamic marker of drug activity^23^. EV may also induce antitumor response of host immune cells, adding to its synergistic benefit when combined with ICIs^4^. This potential immunogenic antitumor response may help explain the higher prevalence and earlier onset of cAEs observed with EV-ICI combination treatment, possibly via a ‘second-hit’ mechanism of immune-mediated cutaneous toxicity^24^. Further molecular validation of these pathways is needed.

Taken together, our findings clarify heterogeneity in prior literature by demonstrating the prognostic value of distinguishing EV-induced cAEs from those attributable to concurrent ICI or other cancer therapies. The timing and granular characterization of distinct morphologies attributable to EV provide additional useful clinical insights and may aid treating physicians in ascribing causality, anticipating toxicity course, and guiding dermatologic toxicity management.

### Limitations

Our study has several limitations. Most notably, its retrospective nature poses a potential for residual confounding despite adjusting for a wide range of covariates to mitigate these effects. Though our sensitivity model accounted for initial EV dose, it did not account for treatment interruption or reduction after EV initiation. Furthermore, attribution of cAEs to EV relied on structured chart review rather than standardized prospective criteria and is therefore subject to observer bias. This is especially relevant for patients not formally evaluated by a dermatologist, particularly in cases of overlapping morphology, timing, and histopathologic features. Third, cAE morphology, severity, and timing characterization was limited by the retrospective availability of clinical descriptions, photographs, and availability of histology. Despite these limitations, the consistent association between EV-induced cAEs and improved survival across various analytic approaches supports the robustness of the findings and highlights the need for future prospective studies.

### Conclusions

In this multi-institutional cohort study, EV-induced cAEs were independently associated with improved survival outcomes, suggesting that cAEs may serve as an early biomarker of therapeutic efficacy among patients treated with EV and their distinct temporal characteristics may help drive treatment decisions for clinicians. Further prospective studies of standardized dermatologic assessment and molecular characterization are needed to better understand the etiology, mitigation strategies, and survival association of EV-induced cAEs.

## Data Availability

All data produced in the present study are available upon reasonable request to the authors

## ACKNOWLEDGEMENT

*Concept and design:* Lee, Karagenova, Fuksman, Semenov

Literature search: Lee, Karagenova

*Abstracting, analysis, or interpretation of data:* Lee, Karagenova, Fuksman, Semenov

*Drafting of the manuscript:* Lee, Karagenova

*Critical revision of the manuscript for important intellectual content:* All authors

*Statistical analysis:* Fuksman, Semenov

*Administrative, technical, or material support:* Reynolds, Semenov

*Supervision:* Semenov

*Data analysis:* Analysis was completed primarily by author Fuksman. Authors Lee, Karagenova, and Semenov had full access to all the data in the study and take responsibility for the integrity of the data and the accuracy of the data analysis.

## Other Acknowledgements

The authors are all incredibly grateful to the patients whose data are enclosed in this manuscript.

