## Supplementary Material for "Enfortumab vedotin-induced cutaneous toxicities and their association with survival in urothelial carcinoma"

**Supplementary Table 1.** Likelihood Scoring Rubric

| | Likelihood Score | | | | |
|---|---|---|---|---|---|
| | 0 | 1 | 2 | 3 | 4 |
| Did the patient have a pre-existing cutaneous eruption before EV initiation?[a] | Yes | No<br>All flares of preexisting conditions (e.g., psoriasis) should not be documented as cAEs. There is a separate section to record flare of pre-existing conditions. | | | |
| Was the cAE more likely due to another etiology (e.g. vancomycin, infection-related)?[b] | N/A | Yes. A different agent or etiology is more likely than EV to cause cAE. | No.<br>Specific likelihood will depend on the questions below. | | |
| Is there documentation the cutaneous eruption is related to EV therapy? | N/A | No | Yes<br>Specific likelihood will depend on the questions below.<br>If the clinician (dermatologist or oncologist) firmly attributes the eruption to EV, it is at least a 3. If they are unsure, it is more likely a 2. | | |
| Is there a medication that is taken concurrently at time of cutaneous eruption? | N/A | N/A | If there is a concurrent medication, but the cutaneous eruption more likely attributed to the concurrent medication | If there is a concurrent medication, but the cutaneous eruption is more likely attributed to EV | If there is no concurrent medication that can lead to cutaneous eruption, the likelihood can be 4 pending answers below. |

**Supplementary Table 1.** Likelihood scoring rubric (continued).

| | Likelihood Score | | | | |
|---|---|---|---|---|---|
| | 0 | 1 | 2 | 3 | 4 |
| Is the diagnosis of EV-induced cAE confirmed by a dermatologist? | N/A | N/A | No | No confirmation by dermatologist. | Yes, the dermatologist confirms the diagnosis. |
| Is the diagnosis of EV-induced cAE confirmed by biopsy? | | | If biopsy results are inconclusive. | | Biopsy confirms EV-induced diagnosis. |

Abbreviations: EV, enfortumab vedotin; cAE, cutaneous adverse event; ICI, immune checkpoint inhibitor.
[a] Likelihood scoring did not apply if no cutaneous eruption occurred after EV initiation.

**Supplementary Methods:**

*Likelihood Scoring*

Cutaneous adverse events (cAEs) were adjudicated by manual chart review by two trained independent reviewers (E.L. and R.K), with resolution of discordant cases by a third reviewer (Y.R.S., a board-certified dermatologist with expertise in oncodermatology). A score of 0 was assigned to a patient who had an eruption prior to initiation of EV, such as a pre-existing diagnosis of psoriasis. A score of 1 (highly unlikely) to 4 (highly likely) that an eruption was attributed to EV was assigned to each patient. Scores were determined by manual review of patient charts and documentation that included clinical photographs, narrative description and assessment by a dermatologist (where patients were referred to dermatology), oncology notes, and timing of an eruption relative to initiation of EV, ICI, or another medication. In cases where a referral to dermatology was not available at the time of an eruption to adjudicate etiology, the reviewers retrospectively analyzed the available documentation to determine likelihood score and morphology. A highly likely (4) score was assigned if there was documentation by a clinician (dermatologist/oncodermatologist) of an EV-related etiology and/or confirmation by histology, along with appropriate timing, lack of competing medications that could contribute to the eruption, and/or other supportive evidence from clinical images and histology. A likely score (3) was assigned if there was a competing medication present, but the eruption was more likely triggered by EV given timing and clinician notes as above. An unlikely (2) score was assigned if there was a competing medication (ICI) that, given timing and clinician notes, was more likely the trigger compared to EV. A highly unlikely (1) score was assigned if an eruption did not coincide with timing of either EV or ICI, and was more likely due to an unrelated drug etiology, such as vancomycin, bactrim, etc. Of note, in cases of uncertainty, scores were assigned conservatively, with lower scores assigned where the above features of an eruption were not definitively supportive of an EV-induced etiology. Patients with a likelihood score of 3 and 4 were categorized as having an EV-induced eruption, and those with scores of 2 and 1 were categorized as non-EV-induced eruptions.

Likelihood scoring methods were adapted from existing literature on cirAE adjudication by Zhang et al, 2023[1], Nguyen et al, 2023[2], Wan et al 2024[3], and Khattab et al 2025[4].

**Supplementary Table 2.** Cancer Treatment Information

| Treatment | No cAE (n=243) | cAE (n=206) | P-value |
|---|---|---|---|
| **Prior ICI** | 102 | 123 | 0.85 |
| **Prior Chemotherapy** | 158 | 120 | 0.145 |
| **Initial EV dose** | | | |
| Full (1.25 mg/kg) | 163 | 162 | 0.00797 |
| Reduced | 80 | 44 | |
| **Total EV infusions** | | | |
| Mean (SD) | 11.7 (10.5) | 16.5 (14.9) | <0.001 |
| Median [Min, Max] | 9.0 [1.00, 55.0] | 14.0 [1.00, 140] | |
| IQR | 13.0 | 13.8 | |

Abbreviations: cAE, cutaneous adverse event; EV, enfortumab vedotin; ICI, immune checkpoint inhibitor; IQR, interquartile range; SD, standard deviation.

**Supplementary Table 3.** Characteristics and Management of cAE by cAE Severity

| | Grade 1 (n=123) | Grade 2 (n=44) | Grade 3 (n=35) | Grade 4 (n=3) | Grade 5 (n=1) | P-value |
|---|---|---|---|---|---|---|
| **Duration of worst cAE (days)** | | | | | | |
| Mean (SD) | 80.4 (110) | 117 (99.2) | 202 (374) | 139 (175) | NA | 0.005 |
| Median [Min, Max] | 41.0 [3.00, 737] | 108 [6.00, 393] | 53.0 [11.0, 1,850] | 65.0 [14.0, 339] | NA | |
| IQR | | | | | NA | |
| **Systemic corticosteroid usage** | | | | | | |
| No | 117 (95.1%) | 33 (75.0%) | 14 (40.0%) | 1 (33.3%) | 1 (100%) | 0.001 |
| Yes | 6 (4.9%) | 11 (25.0%) | 21 (60.0%) | 2 (66.7%) | 0 (0%) | |
| **Dupilumab usage** | | | | | | |
| No | 123 (100%) | 39 (88.6%) | 31 (88.6%) | 3 (100%) | 1 (100%) | 0.004 |
| Yes | 0 (0%) | 5 (11.4%) | 4 (11.4%) | 0 (0%) | 0 (0%) | |
| **Topical corticosteroid usage** | | | | | | |
| No | 28 (22.8%) | 7 (15.9%) | 5 (14.3%) | 0 (0%) | 1 (100%) | 0.163 |
| Yes | 95 (77.2%) | 37 (84.1%) | 30 (85.7%) | 3 (100%) | 0 (0%) | |
| **Cumulative prednisone-equivalent dose (mg)** | | | | | | |
| Mean (SD) | 521 (586) | 649 (554) | 1,180 (1,590) | 1260 (328) | NA | 0.687 |
| Median [Min, Max] | 353 [105, 1680] | 400 [155, 1500] | 440 [27, 6240] | 1260 [1030, 1500] | NA | |
| IQR | 280 | 937 | 1,280 | 232 | NA | |
| **Duration of systemic corticosteroid (days)** | | | | | | |
| Mean (SD) | 64.7 (46.8) | 60.7 (59.9) | 67.9 (91.2) | 74.0 (58.0) | NA | 0.651 |
| Median [Min, Max] | 55.5 [21.0, 125] | 49.0 [5.00, 192] | 24.0 [0, 392] | 74.0 [33.0, 115] | NA | |
| IQR | 78.0 | 81.0 | 86.0 | 41.0 | NA | |
| **Average daily prednisone-equivalent dose (mg)** | | | | | | 0.191 |
| Mean (SD) | 10.2 (8.44) | 22.8 (20.1) | 32.2 (38.0) | 22.1 (12.9) | NA | |
| Median [Min, Max] | 7.69 [1.24, 20.9] | 20.0 [1.96, 66.8] | 20.6 [3.39, 150] | 22.1 [13.0, 31.2] | NA | |
| IQR | 13.4 | 27.4 | 17.9 | 9.12 | NA | |

Abbreviations: cAE, cutaneous adverse event; IQR, interquartile range; SD, standard deviation.

**Supplementary Figure 1.** Distribution of Median Time to Worst Cutaneous Adverse Event

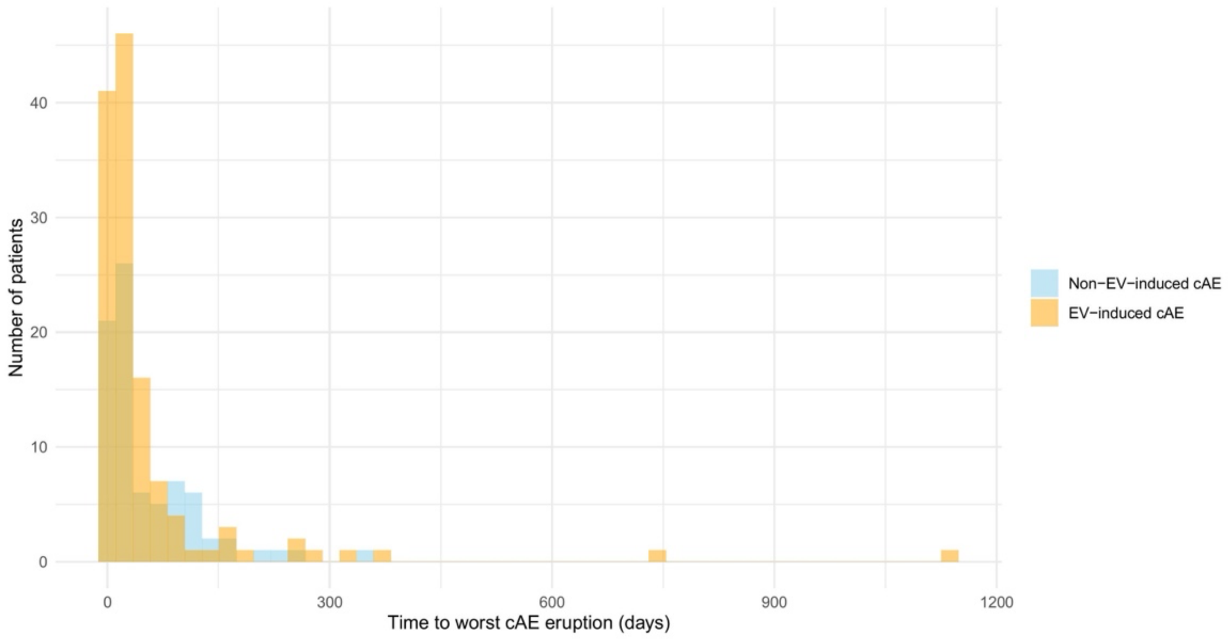

**Supplementary Table 4.** Cutaneous Adverse Event Morphology Distribution Stratified by Treatment Group and Likelihood Scoring[a]

| | EV-only | | EV+ICI | | EV+other | |
|---|---|---|---|---|---|---|
| **Cutaneous Adverse Event** | **Non-EV-induced cAE (%)[b] (n=10)** | **EV-induced cAE (%)[b] (n=78)** | **Non-EV-induced cAE (%)[b] (n=93)** | **EV-induced cAE (%)[b] (n=89)** | **Non-EV-induced cAE (%)[b] (n=7)** | **EV-induced cAE (%)[b] (n=16)** |
| Pruritus | 3 (30.0) | 20 (25.7) | 30 (32.3) | 25 (28.1) | 2 (28.6) | 5 (31.3) |
| Rash and Other Unspecified Eruption | | | | | | |
| Rash (unspecified) | 4 (40.0) | 15 (19.2) | 26 (28.0) | 12 (13.5) | 3 (42.9) | 3 (18.8) |
| Desquamative dermatitis | 0 (0) | 4 (5.1) | 2 (2.2) | 7 (7.9) | 0 (0) | 0 (0) |
| Morbilliform Dermatitis | 1 (10.0) | 18 (23.1) | 19 (20.4) | 16 (18.0) | 0 (0) | 4 (25.0) |
| Bullous Dermatitis | 0 (0) | 8 (10.3) | 4 (4.3) | 5 (5.6) | 0 (0) | 0 (0) |
| Other[c] | 0 (0) | 6 (7.7) | 2 (2.2) | 2 (2.2) | 1 (14.3) | 0 (0) |
| Xerosis Cutis | 1 (10.0) | 3 (3.8) | 3 (3.3) | 2 (2.2) | 0 (0) | 0 (0) |
| SDRIFE | 0 (0) | 1 (1.3) | 5 (5.4) | 0 (0) | 0 (0) | 1 (6.3) |
| Eczematous | | | | | | |
| Eczematous Dermatitis | 0 (0) | 0 (0) | 3 (3.3) | 4 (4.5) | 0 (0) | 1 (6.3) |
| Eczematous And Other | 0 (0) | 0 (0) | 1 (1.1) | 0 (0) | 0 (0) | 0 (0) |
| Lichenoid | | | | | | |
| Lichenoid Dermatitis | 1 (10.0) | 2 (2.6) | 2 (2.2) | 2 (2.2) | 0 (0) | 0 (0) |
| Lichenoid And Other | 0 (0) | 0 (0) | 0 (0) | 3 (3.4) | 0 (0) | 0 (0) |
| Alopecia | 0 (0) | 1 (1.3) | 0 (0) | 0 (0) | 1 (14.3) | 1 (6.3) |
| Mucositis | 0 (0) | 1 (1.3) | 1 (1.1) | 0 (0) | 0 (0) | 1 (6.3) |
| Psoriasiform Dermatitis | 0 (0) | 0 (0) | 0 (0) | 2 (2.2) | 0 (0) | 0 (0) |
| Toxic Epidermal Necrolysis | 0 (0) | 0 (0) | 0 (0) | 2 (2.2) | 0 (0) | 0 (0) |
| Lichen Simplex Chronicus | 0 (0) | 0 (0) | 0 (0) | 1 (1.1) | 0 (0) | 0 (0) |
| Pyoderma Gangrenosum | 0 (0) | 0 (0) | 0 (0) | 1 (1.1) | 0 (0) | 0 (0) |

Abbreviations: SDRIFE, Symmetrical Drug-Related Intertriginous and Flexural Exanthema.

[a]Likelihood scores of 3 or 4 were considered as EV-induced in this study, scores of 0-2 considered as non-EV- induced associated cAEs, and no likelihood scoring was not applied if no cAE developed.

[b]Percentages refer to the number of patients with associated cAE. Percentages do not sum to 100 because some patients may have had multiple cAEs.

[c]Other includes skin ulceration, bruising, petechial rash, papular follicular rash, erythroderma, cellulitis, interface dermatitis.

**Supplementary Table 5.** Median Survival Times Stratified by EV-induced and non-EV-induced cAE

| **Treatment Group** | **Median PFS (months)** | **Median OS (months)** |
|---|---|---|
| **EV-only** | 5.2 | 10.9 |
| no cAE | 3.7 | 7.9 |
| non-EV-induced cAE | 6.8 | 13.1 |
| EV-induced cAE | 6.9 | 19.7 |
| **EV+ICI** | 10.6 | 21.0 |
| no cAE | 5.7 | 9.8 |
| non-EV-induced cAE | 18.5 | N/A |
| EV-induced cAE | 26.3 | N/A |
| **EV+other** | 11.3 | 16.7 |
| no cAE | 10.8 | 14.7 |
| non-EV-induced cAE | 5.7 | 13.0 |
| EV-induced cAE | 13.3 | 25.6 |

Abbreviations: cAE, cutaneous adverse event; EV, enfortumab vedotin; ICI, immune checkpoint inhibitor; OS, overall survival; PFS, progression-free survival.

**Supplementary Figure 2.** Kaplan-Meier Estimates of Progression-Free Survival (PFS) (panel A) and Overall Survival (OS) (panel B) in EV+other treatment group stratified by EV-induced, non-EV-induced cAE, or no cAE

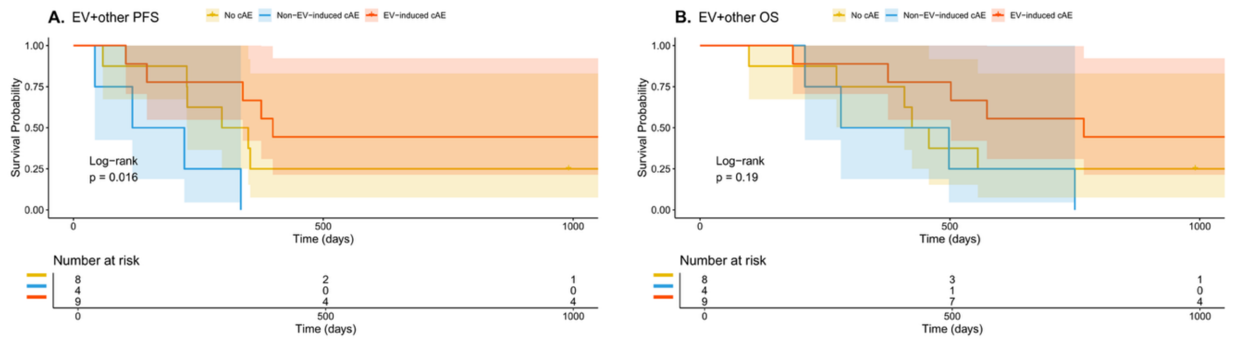


EV+other cohort includes patients treated with combination of EV and sacituzumab govitecan, EV and ALX148 (anti-CD47), and EV and GEN-009 vaccine.

**Supplementary Figure 3.** Hazard Rate for EV-induced cAE Progression-Free Survival (PFS) (panel A) and Overall Survival (OS) (panel B) and for Early Onset EV-induced cAE for PFS (panel C) and OS (panel D) across Different Landmark Times

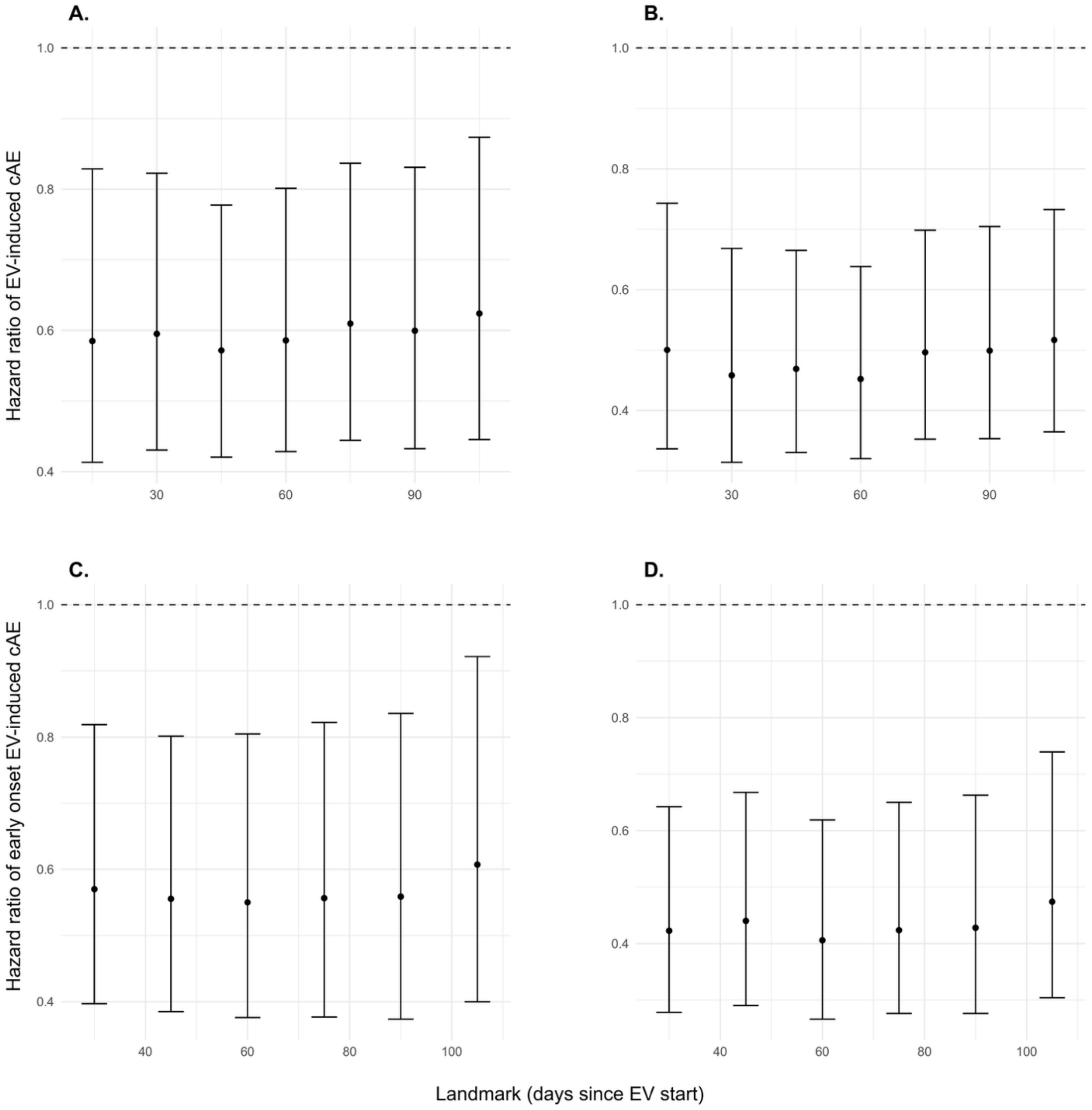

**Supplementary Figure 4.** Hazard Rate for EV-induced cAE Progression-Free Survival (PFS) (panel A) and Overall Survival (OS) (panel B) and for Early Onset EV-induced cAE for PFS (panel C) and OS (panel D) across Different Landmark Times, adjusting for ECOG-PS, initial EV dose, prior ICI and prior chemotherapy exposure.

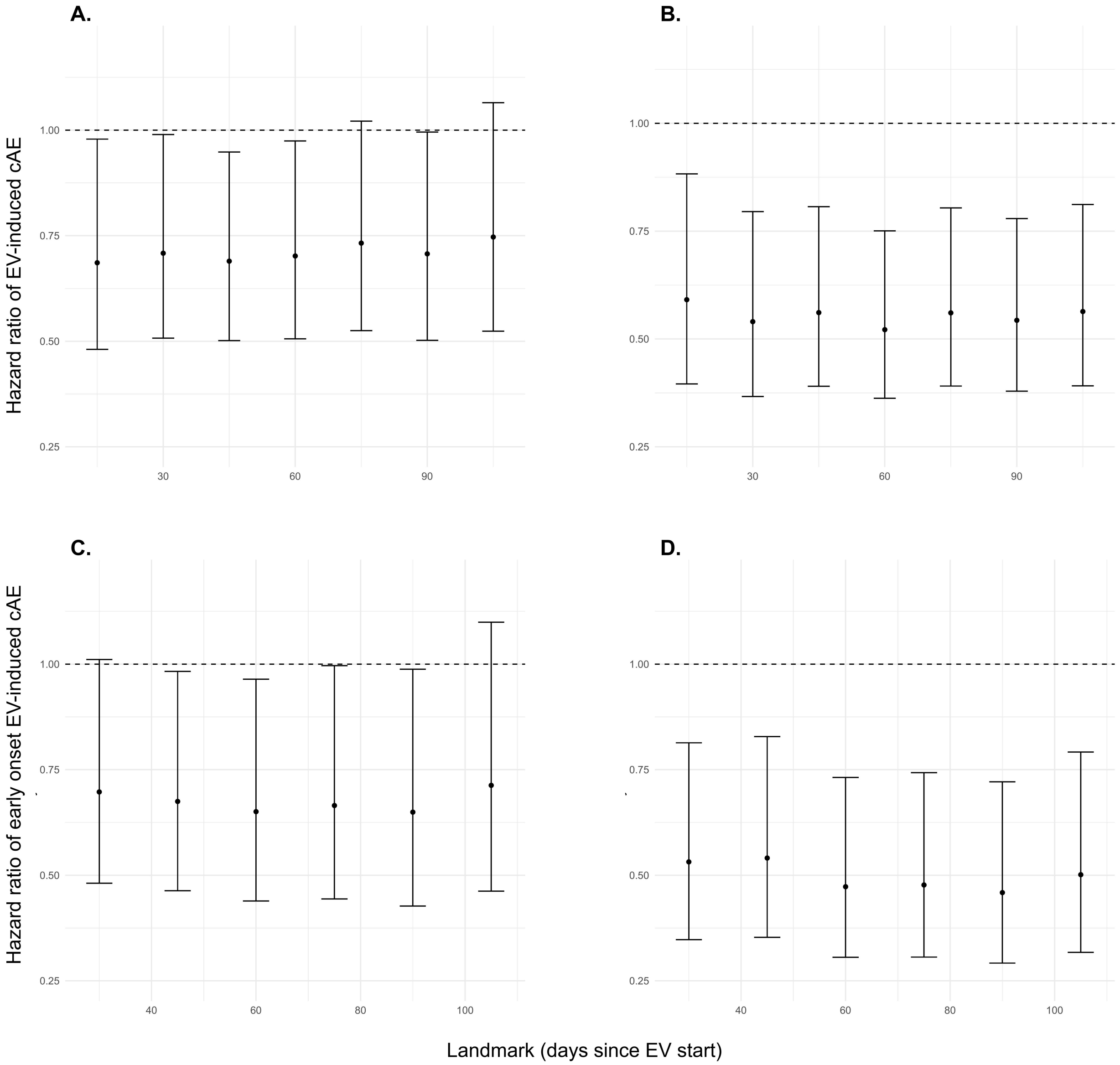


Abbreviations: cAE, cutaneous adverse event; ECOG-PS, Eastern Cooperative Oncology Group Performance Status; EV, enfortumab vedotin; ICI, immune checkpoint inhibitor.

**Supplementary Table 6.** Most Common Non-Cutaneous Adverse Events Stratified by Treatment Group and Likelihood Scoring[a]

| Non-Cutaneous Adverse Event | EV-only (%)[b] (n=159) | EV+ICI (%)[b] (n=251) | EV+other (%)[b] (n=30) | Total (%)[c] (n=449) | EV-induced cAE (%)[b] (n=158) | Non-EV-induced cAE (%)[b] (n=89) | No cAE (%) (n=193)[b] |
|---|---|---|---|---|---|---|---|
| Neuropathy | 65 (40.9) | 93 (37.1) | 12 (40.0) | 170 (37.9) | 66 (41.8) | 39 (43.8) | 65 (33.7) |
| Fatigue | 33 (20.8) | 48 (19.1) | 4 (13.3) | 85 (18.9) | 32 (20.3) | 12 (13.5) | 41 (21.2) |
| Diarrhea | 14 (8.8) | 20 (8.0) | 6 (20.0) | 40 (8.9) | 15 (9.5) | 6 (6.7) | 19 (9.8) |
| Failure to Thrive | 6 (3.8) | 16 (6.4) | 0 (0) | 22 (3.9) | 4 (2.5) | 3 (3.4) | 15 (7.8) |
| Anorexia | 8 (5.0) | 11 (4.4) | 0 (0) | 19 (4.2) | 4 (2.5) | 1 (1.1) | 13 (6.7) |
| Nausea | 7 (4.4) | 9 (3.6) | 2 (6.7) | 18 (4.0) | 5 (3.2) | 5 (3.2) | 11 (5.7) |
| Hyperglycemia | 3 (1.9) | 11 (4.4) | 0 (0) | 14 (3.1) | 5 (3.2) | 5 (5.6) | 4 (2.1) |
| Ocular Irritation | 4 (2.5) | 5 (2.0) | 3 (10.0) | 12 (2.7) | 6 (3.8) | 5 (5.6) | 1 (0.5) |
| Hepatitis | 3 (1.9) | 7 (2.8) | 1 (3.3) | 11 (2.4) | 5 (3.2) | 4 (4.5) | 2 (1.0) |
| Appetite Loss | 3 (1.9) | 7 (2.8) | 0 (0) | 10 (2.2) | 3 (1.8) | 2 (2.2) | 5 (2.5) |
| Dysgeusia | 3 (1.9) | 6 (2.4) | 0 (0) | 9 (2.0) | 2 (1.3) | 4 (4.5) | 3 (1.6) |
| Anemia | 4 (2.5) | 1 (0.4) | 0 (0) | 5 (1.1) | 1 (0.6) | 0 (0) | 4 (2.1) |

Abbreviations: cAE, cutaneous adverse event; EV, enfortumab vedotin; ICI, immune checkpoint inhibitor.
[a]Likelihood scores of 3 or 4 were considered as EV-induced in this study, scores of 0-2 considered as non-EV-induced associated cAEs, and no likelihood scoring was not applied if no cAE developed.
[b]Percentages represent proportion of non-cutaneous adverse event in their respective group. Number in each group reflects total number of non-cutaneous adverse events, which accounts for why they are not equal to total number of patients in each group type.
[c]Total percentage represents proportion of the non-cutaneous adverse event in the total cohort.
